# A Systematic Review of Interventions for Prevention and Treatment of Post-Traumatic Stress Disorder Following Childbirth

**DOI:** 10.1101/2023.08.17.23294230

**Authors:** Sharon Dekel, Joanna E. Papadakis, Beatrice Quagliarini, Kathleen M. Jagodnik, Rasvitha Nandru

**Author notes:** **Corresponding Author:** Sharon Dekel, Ph.D. Department of Psychiatry, Harvard Medical School & Massachusetts General, Hospital, Boston, USA.

## Abstract

**Objective:** Postpartum women can develop post-traumatic stress disorder (PTSD) in response to complicated, traumatic childbirth; prevalence of these events remains high in the U.S. Currently, there is no recommended treatment approach in routine peripartum care for preventing maternal childbirth-related PTSD (CB-PTSD) and lessening its severity. Here, we provide a systematic review of available clinical trials testing interventions for the prevention and indication of CB-PTSD.

**Data Sources:** We conducted a systematic review of PsycInfo, PsycArticles, PubMed (MEDLINE), ClinicalTrials.gov, CINAHL, ProQuest, Sociological Abstracts, Google Scholar, Embase, Web of Science, ScienceDirect, and Scopus through December 2022 to identify clinical trials involving CB-PTSD prevention and treatment.

**Study Eligibility Criteria:** Trials were included if they were interventional, evaluated CB-PTSD preventive strategies or treatments, and reported outcomes assessing CB-PTSD symptoms. Duplicate studies, case reports, protocols, active clinical trials, and studies of CB-PTSD following stillbirth were excluded.

**Study Appraisal and Synthesis Methods:** Two independent coders evaluated trials using a modified Downs and Black methodological quality assessment checklist. Sample characteristics and related intervention information were extracted via an Excel-based form.

**Results:** A total of 33 studies, including 25 randomized controlled trials (RCTs) and 8 non-RCTs, were included. Trial quality ranged from Poor to Excellent. Trials tested psychological therapies most often delivered as secondary prevention against CB-PTSD onset (n=21); some examined primary (n=3) and tertiary (n=9) therapies. Positive treatment effects were found for early interventions employing conventional trauma-focused therapies, psychological counseling, and mother-infant dyadic focused strategies. Therapies’ utility to aid women with severe acute traumatic stress symptoms or reduce incidence of CB-PTSD diagnosis is unclear, as is whether they are effective as tertiary intervention. Educational birth plan-focused interventions during pregnancy may improve maternal health outcomes, but studies remain scarce.

**Conclusions:** An array of early psychological therapies delivered in response to traumatic childbirth, rather than universally, in the first postpartum days and weeks, may potentially buffer CB-PTSD development. Rather than one treatment being suitable for all, effective therapy should consider individual-specific factors. As additional RCTs generate critical information and guide recommendations for first-line preventive treatments for CB-PTSD, the psychiatric consequences associated with traumatic childbirth could be lessened.

**Disclosure Statement:** The authors report no conflict of interest.

**Financial Support and Roles of Funding Sources:** Dr. Sharon Dekel was supported by grants from the National Institute of Child Health and Human Development (R01HD108619, R21HD100817, and R21HD109546) and an ISF award from the Massachusetts General Hospital Executive Committee on Research. Dr. Kathleen Jagodnik was supported by a Mortimer B. Zuckerman STEM Leadership Program Postdoctoral Fellowship. Ms. Joanna Papadakis was supported by a grant through the Menschel Cornell Commitment Public Service Internship at Cornell University. None of the funding organizations had a role in designing, conducting, or reporting this work.

**Information for Systematic Review:**

∼ (i) Date of PROSPERO Registration: 07-12-2021
∼ (ii) Registration Number: CRD42020207086

## Introduction

Childbirth is a profound experience often entailing extreme physical and psychological stress. Among delivering women, an estimated 1/3 experience highly stressful and potentially traumatic birth,^1–3^ and ∼60,000 women in the U.S. each year experience severe maternal morbidity (SMM).^4^ SMM rates in the U.S. are among the highest in Western countries^5–7^ and steadily continue to increase.^5, 8–10^

Complicated deliveries may undermine maternal psychological welfare. Post-traumatic stress disorder (PTSD) is the formal psychiatric disorder resulting from exposure to an event involving life-threat or physical harm and associated psychological symptoms that do not resolve naturally over time.^11^ Existing research supports the validity of PTSD following childbirth, or childbirth-related PTSD (CB-PTSD).^12^ The prevalence of this condition is estimated at 5-6% of all postpartum women;^3, 13, 14^ this translates nationally to 240,000 affected American women each year. In complicated deliveries, 18.5% to 41.2% of women^14–16^ report CB-PTSD symptoms. Black and Latinx women are nearly three times more likely to endorse a childbirth-related traumatic stress response.^17^

Although highly co-morbid with peripartum depression,^18–20^ CB-PTSD is a distinct condition largely consistent with the formal symptom constellation of PTSD.^21^ Exposure to a traumatic childbirth can result in childbirth-related involuntary intrusion symptoms such as flashbacks and nightmares; attempts to avoid reminders of childbirth; negative alterations in cognitions and mood, and marked arousal and reactivity manifested in irritability, sleep and concentration problems, hypervigilance, and other symptoms.^22–24^

When left untreated, CB-PTSD can impair maternal functioning during the important postpartum period. Women with CB-PTSD may exhibit reduced maternal affection, bonding, and sensitive behavior toward their infant,^25–28^ which may increase the risk for social and emotional developmental problems in the infant.^27, 29^ Available research suggests that maternal CB-PTSD associates with infant behavioral problems, as well as sleep and feeding problems, including less favorable breastfeeding outcomes.^29^ Untreated CB-PTSD can also result in avoidance of partner intimacy and disincentivize future pregnancies.^30–32^

CB-PTSD has unique attributes that support the potential for early intervention and even prevention. PTSD symptoms begin after a specified external traumatic event (here, childbirth) and appear in the first days following exposure,^18, 33, 34^ suggesting that CB-PTSD follows a clear onset. Theoretical models of non-childbirth PTSD pathogenesis suggest that beyond pre-existing vulnerabilities, biological and psychological mechanisms underlie an individual’s immediate response to a traumatic event that could be targeted by interventions to buffer and avert the PTSD trajectory.^35–38^ Consequently, early interventions could produce favorable outcomes. Unlike other forms of trauma, childbirth is a relatively time-defined event for which women often stay in the hospital after parturition. This suggests an important opportunity to identify and treat women before they develop the full traumatic stress syndrome.

Presently, there is a critical gap in knowledge to inform recommendations to prevent and treat CB-PTSD. Early review studies on this topic used the limited number of available clinical trials, preventing firm conclusions about the utility of psychological debriefing and individual counseling therapies.^39–41^ In recent years, 6 systematic reviews have been performed that focused mostly on early interventions; they included 45 trials published up to 2022.^42–47^ The reviews concluded that early-administered trauma-focused interventions that work through exposure and reprocessing of the traumatic memory and related cognitions appear helpful for alleviating symptoms of CB-PTSD in the short term, but that more studies were warranted to establish clinical recommendations.

## Objectives

We provide a comprehensive systematic review of randomized and non-randomized controlled clinical trials for preventing CB-PTSD onset or reducing symptoms severity in affected women. We used a quantitative rating system to evaluate the published trials’ quality.^48^ To the best of our knowledge, this approach has not previously been implemented. We reviewed the potential benefits of primary, secondary, and tertiary prevention approaches for CB-PTSD to provide insight into which therapies are most promising, what the optimal timing for intervention may be, and which populations will benefit most from these interventions. Publication dates range from December 1998 to December 2022.

## Methods

### a. Eligibility Criteria, Information Sources and Search Strategy

To be included in this review, studies were independently evaluated based on the following inclusion criteria: a) interventional study; b) indication of CB-PTSD prevention or treatment; and c) inclusion of outcome measure(s) assessing CB-PTSD symptoms or diagnosis. Duplicate studies, case reports, study protocols, active clinical trials, and studies involving mothers who exclusively experienced stillbirth were excluded.

This systematic review was conducted according to PRISMA guidelines,^49^ and our protocol is registered on PROSPERO (CRD42020207086). Our search strategy targeted all published studies measuring CB-PTSD or its symptoms as a primary treatment outcome. Articles published through December 2022 were included from the following databases: PsycInfo, PsycArticles, PubMed (MEDLINE), ClinicalTrials.gov, CINAHL, ProQuest, Sociological Abstracts, Google Scholar, Embase, Web of Science, ScienceDirect, and Scopus. The search criteria employed any combination of these keywords: "Postpartum OR postnatal OR childbirth PTSD" OR "traumatic childbirth" OR "childbirth induced post-traumatic stress" AND "treatment OR intervention OR therapy OR prevention”. Published reviews on CB-PTSD therapies served as additional resources.

### b. Study Selection

A total of 33 studies published from December 1998 to December 2022 met inclusion criteria and were reviewed. This selection followed the PRISMA workflow process;^49^ for more information regarding study selection, see Figure 1.

**Figure 1.**
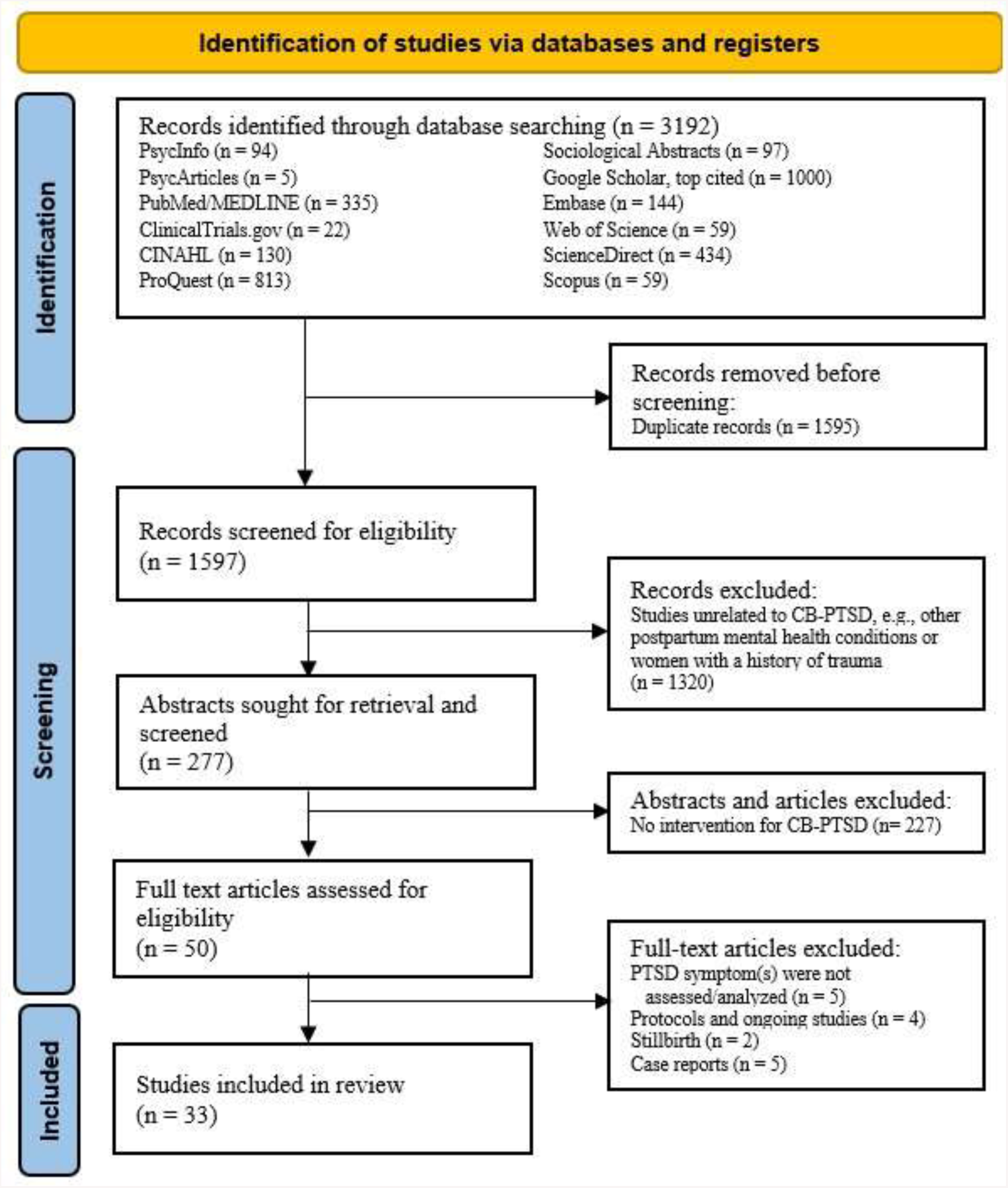
PRISMA flow diagram detailing the source selection process of both randomized and non-randomized clinical studies targeting childbirth-related post-traumatic stress disorder (CB-PTSD) in at-risk and universal samples.

### c. Data Extraction

Two reviewers (J.P. and R.N.) extracted data using an Excel-based form. For each study, the reviewers collected: sample characteristics, treatment type (prevention/treatment) and modality, intervention frequency/duration, primary outcome measures, and outcome time points (immediate, moderate, and long-term). We report treatment effects on CB-PTSD symptoms and related conditions. Details are presented in Table 1.

**Table 1.** Descriptions of clinical trials for interventions to prevent or treat childbirth-related maternal PTSD.

### d. Assessment of Risk of Bias, and Quality Assessment

We adopted the well-validated, commonly used Downs and Black checklist^48^ that is recommended for evaluating the quality of randomized and non-randomized healthcare interventions. This 27-item checklist offers a quantitative rating scale that is a composite measurement of external validity, internal validity/confounding bias, and statistical power. Individual items are scored on an integer scale of 0 to 1, for a total score of 28 (with item 5 scored 0-2). A study’s overall quality score is calculated using the checklist’s assigned point system, with higher score indicating higher study quality. In this review, to better identify high-quality trials, items 20, 21, and 27 ("Were the main outcome measures used accurate?”; “Were the patients in different intervention groups or were the cases and controls recruited from the same population?”; “Were study subjects randomized to intervention groups?”, respectively) were scored on a 0-2 scale, as done in previous studies,^50–52^ with a maximum total score of 31. Modified quality score ranges were specified as: Excellent (29-31); Good (22-28); Fair (17-21); Poor (≤16).

The two reviewers independently scored all 33 studies. Inter-rater reliability was high (91%), and any discrepancy in scores was discussed until 100% agreement was achieved. The quality scores and adopted checklist are presented in Table 1 and Appendix A, respectively.

## Results

### a. Study Selection

In this review of 33 studies, 25 were randomized controlled trials (RCTs) and 8 were non-RCTs. Among all trials, 3 tested primary preventive interventions delivered during pregnancy; 19 tested secondary preventions in which treatment was provided after childbirth but not later than 1 month postpartum, i.e., before a DSM PTSD diagnosis can be confirmed;^11^ and 11 trials involved tertiary prevention delivered more than 1 month postpartum, in cases with confirmed or probable CB-PTSD.

Of the 33 studies, 31 entailed psychologically oriented therapies. These included trauma-focused structured and non-structured interventions (n=19), i.e., psychological debriefing, crisis intervention, Trauma-Focused Cognitive Behavioral Therapy (TF-CBT), Eye Movement Desensitization and Reprocessing (EMDR), and Trauma-Focused Expressive Writing (TF-EW); mother-infant dyad therapies (n=3); psychological counseling (n=5); and other psychological approaches, i.e., visual biofeedback (n=1) and visual spatial cognitive task (n=2). The remaining 3 trials were educational interventions (Table 1).

In most trials, treatment response was determined using validated patient self-administered questionnaires to measure endorsement of CB-PTSD symptoms (n=30). Clinician evaluation to determine CB-PTSD endorsement was performed in 3 trials. Assessment of sustained treatment outcomes usually targeted the first months following the intervention (n=27, ≥1.5 months, ≤6 months); longer effects (≥12 months post-intervention) were measured in 6 trials.

### b. Study Characteristics and Risk of Bias Results

Detailed information on the study characteristics and quality assessment results are presented in Table 1.

### c. Synthesis of Results

#### Psychologically Oriented

#### Debriefing or Trauma-Focused Psychological Therapies (TFPT)

Psychological debriefing in the postpartum is usually performed as an early intervention via midwife-led dialogue involving delivery-related emotions.^44, 53, 54^ When treating CB-PTSD, the stressful aspects of the childbirth experience are addressed. Debriefing was tested in 4 RCTs as early secondary prevention and 1 NRCT as later (tertiary) prevention. Quality scores ranged from Fair to Good (17-28).

Overall, although women generally consider debriefing of value, no evidence supports the psychological benefit of midwife-led postpartum debriefing following healthy^55, 56^ or complicated^57, 58^ deliveries. A single structured 15-60 minute debriefing session within 72 hours post-delivery vs. treatment as usual (TAU) was not associated with reduction in traumatic stress, depression,^55, 56^ anxiety, or parenting distress,^55^ nor the proportion of incidences meeting CB-PTSD diagnosis.^56^ A sub-group of women receiving debriefing who experienced more medical interventions had *more* negative perceptions of childbirth than controls.^55^ Similarly, no sustained benefits were documented when debriefing was offered to women following complicated, traumatic deliveries, compared with cognitive behavioral therapy (CBT) and/or TAU.^57, 58^

Debriefing offered as a later treatment for women possibly affected by CB-PTSD symptoms is associated with positive outcomes; however, evidence is derived from a single NRCT. Compared with TAU (no debriefing), a single (60-90 minutes) debriefing session delivered ∼16 weeks postpartum upon maternal request/referral reduced CB-PTSD symptoms and negative appraisals of childbirth, but not depression, in a sample of 80 women who met DSM Criterion A.^59^

#### Crisis Intervention

Trauma-Focused (TF) crisis intervention entails providing information about the impact of stress, identifying relevant resources, and learning relaxation techniques and coping strategies in the aftermath of trauma.^60^ A single NRCT tested TF intervention as secondary prevention for CB-PTSD; quality score was Fair (19).^60^

An early single-session TF crisis intervention performed in the neonatal intensive care unit (NICU) in women experiencing premature delivery and therefore at elevated risk for CB-PTSD showed short-term benefits.^60^ The intervention was tested in an NRCT of 50 mothers of premature infants and was coupled with brief psychological aid and intense support during critical times. At time of hospital discharge, mothers receiving the intervention compared with TAU (can receive hospital minister counseling) had fewer overall CB-PTSD symptoms and fewer intrusion, avoidance, and hyperarousal symptoms.

#### Trauma-Focused Cognitive Behavioral Therapy

Trauma-Focused Cognitive Behavioral Therapy (TF-CBT) for CB-PTSD involves a manualized protocol in which cognitive distortions regarding the traumatic childbirth and related stressors are challenged to reorient adaptive thoughts and behaviors.^23, 42, 61^ TF-CBT was tested in 1 RCT and 1 NRCT as secondary prevention,^62–64^ and in 2 RCTs as tertiary prevention.^65, 66^ Quality scores ranged from Fair to Good (20-27).

Early TF-CBT delivered during premature infant’s hospitalization in women at risk for CB-PTSD shows benefits.^62, 63^ The therapy involves cognitive restructuring, muscle relaxation, construction of a narrative of the traumatic childbirth and NICU experience, psychoeducation, and infant redefinition.^62, 63^ Consecutive (6, ∼50 minutes) TF-CBT one-on-one sessions delivered 1-2 weeks following childbirth, vs. standard care, in an RCT of 105 women with clinically significant acute stress, yielded fewer CB-PTSD and postpartum depression (PPD) symptoms; positive treatment effects were sustained 6 months after childbirth.^62, 63^ Similarly, consecutive (6, 90 minutes) TF-CBT group sessions in the NICU in 19 women (no controls) was associated with improved PPD, CB-PTSD, and anxiety symptoms, at 6 weeks and 6 months post-intervention, respectively.^64^

Findings are mixed for TF-CBT sessions focused on exposure and cognitive restructuring delivered in the months and years postpartum to affected women.^65, 66^ A series of 6-8 consecutive TF-CBT internet sessions delivered 2-4 months following medically complicated delivery was not associated with better long-term outcomes vs. TAU (conventional support) in an RCT (N = 266).^65^ Improvement in CB-PTSD and PPD symptoms, and reported quality of life, assessed 1-year post-treatment were observed in both study conditions. In contrast, consecutive (8 weekly) sessions delivered ∼2.8 years postpartum in women with provisional CB-PTSD^66^ were associated with improvement in CB-PTSD symptoms and reported quality of life compared with delayed (post 5 months) treatment in an RCT of 56 women, although improvement in anxiety and PPD symptoms was observed in both treatment conditions.

#### Eye Movement Desensitization and Reprocessing (EMDR)

Trauma-Focused Eye Movement Desensitization and Reprocessing (TF-EMDR) for CB-PTSD involves a standardized protocol in which women are instructed to focus briefly on their traumatic memories of childbirth while receiving bilateral eye stimulation to reprocess and alleviate childbirth-related traumatic stress.^44^ EMDR was tested in 1 RCT as secondary prevention^67^ and 2 NRCTs as tertiary prevention.^68, 69^ Quality scores ranged from Poor to Good (13-25).

An early TF-EMDR intervention delivered during maternity hospitalization stay can reduce CB-PTSD symptoms in postpartum women at high risk for CB-PTSD, endorsing acute traumatic stress. A single (90-minute) session delivered within 72 hours post-delivery vs. TAU (standard psychological supportive therapy) in women with childbirth-related traumatic stress (N = 37) yielded significant reduction of CB-PTSD symptoms and subjective distress regarding recent and future deliveries, assessed 6 weeks and 3 months later.^67^ However, no group differences were found in the prevalence of CB-PTSD diagnosis post-treatment, and improvement in mother-infant bonding and PPD symptoms was noted in both conditions. In consecutive EMDR sessions in the months and years following childbirth in affected women endorsing CB-PTSD, positive outcomes are reported;^68, 69^ it should be noted that the findings are derived from two NRCTs without control groups.

#### Trauma-Focused Expressive Writing

Trauma-Focused Expressive Writing (TF-EW) for CB-PTSD involves constructing a narrative about childbirth through writing with a focus on describing related thoughts and feelings.^42, 70^ This is intended to facilitate reprocessing of the birth experience and enhance meaning making.^71^ TF-EW was tested in a total of 6 RCTs including 4 trials as secondary prevention and 2 trials as tertiary prevention. Quality scores ranged from Fair to Good (20-27).

TF-EW delivered in the very first days following uncomplicated pregnancies shows benefits. A single (10-15 minute) EW session about childbirth, compared with no writing, performed 48 hours postpartum in samples of 64(72) and 242(73) women, was associated with fewer hyperarousal and avoidance symptoms post-intervention. Sustained positive treatment effects (for hyperarousal) were observed at 2 months^72, 73^ and 1 year^73^ following childbirth. Likewise, a single (∼20 minute) TF-EW session about childbirth, compared with writing about daily events, performed ∼96 hours post-delivery (N = 176), was associated at 3 months post-treatment with positive effects in reducing depressive and PTSD (hyperarousal and avoidance) symptoms.^74^ Immediate treatment effects were observed for depression.

Consecutive TF-EW sessions can benefit postpartum women who are at risk of CB-PTSD. In a non-selective sample of 113 women, early TF-EW (2 sessions in a single day) about childbirth delivered in the first postpartum days, compared with neutral writing, produced greater reduction in CB-PTSD (avoidance and hyperarousal) symptoms, and depressive symptoms 3 months later, especially for women with relatively higher stress at baseline.^75^ Likewise, in a high-risk sample of 67 women, TF-EW (3, 15-minute sessions) in the months following prematurity, focused on the childbirth and infant’s hospitalization, had positive outcomes. Compared with TAU (standard postpartum care), EW was associated with improvements in post-traumatic stress and depressive symptoms, and overall mental health status, 1 month following intervention. Treatment effects (for depression) were sustained 3 months later.^76^ Similarly, TF-EW (4, 30-minute sessions) post-discharge vs. waiting-list, in a sample of 38 postpartum women following premature delivery, was associated with improvement in post-traumatic stress 1 month post-intervention.^77^

#### Psychological Counseling

Psychological counseling for CB-PTSD in postpartum women usually entails semi-structured midwife-led intervention emphasizing the therapeutic relationship, acceptance of childbirth experiences, expression of emotions, social support, problem solving,^39, 78, 79^ and discussion of baby care-related issues.^80^ Psychological counseling was tested in 4 RCTs as secondary prevention, and 1 RCT as tertiary prevention. Quality scores ranged from Fair to Good (18-26).

One-on-one midwife-led psychological counseling in which the core intervention is conducted as a single session in the postpartum unit for women who experience traumatic childbirth shows positive effects.^81, 82^ Two studies of 90 and 103 postpartum women, respectively, who experienced birth trauma and thus met DSM Criterion A, tested a single (40-60 minutes) counseling session within 72 hours post-delivery coupled with a phone session (40-60 minutes) 4-6 weeks postpartum vs. TAU.^81, 82^ Counseling was associated with reduction in CB-PTSD and PPD symptoms,^81, 82^ less self-blame, and greater confidence about future pregnancies 3 months later,^82^ although not reducing incidences of CB-PTSD diagnosis.^82^

A more intense early counseling therapy entailing consecutive sessions delivered in the postpartum unit and subsequently during postpartum weeks to women who experienced traumatic childbirth also showed benefits.^80, 83^ A single one-on-one (45-60 minutes) counseling session delivered 24-48 hours post-delivery followed by a 45-90-minute session during postpartum care visit at 10-15 days, and a brief (15-20 minutes) counseling session via phone 4-6 weeks after delivery vs. TAU (routine post-partum care), were tested in a sample of 166 postpartum women meeting PTSD DSM Criterion A.^80^ Counseling sessions were associated with reduced CB-PTSD and PPD symptoms, and improved maternal-infant bonding at 2 months postpartum.^80^ Likewise, consecutive (2, 45-60 minutes) counseling sessions about the implications and consequences of an emergency Cesarean section, delivered before hospital discharge, and 2 additional sessions performed 2-3 weeks postpartum vs. TAU were tested in 99 postpartum women.^83^ Counseling was associated with fewer CB-PTSD symptoms, less general mental distress, and more positive appraisals of recent childbirth at 1 month postpartum, with effects sustained 6 months postpartum. However, the treatment was insufficient for women with substantial post-traumatic stress reactions or general distress.^83^

In contrast, intervention of later postpartum counseling group-format intervention sessions in months following traumatic childbirth does not appear promising. Consecutive (2, 60 minutes) sessions vs. TAU in 162 women who had emergency Cesarean section did not reduce level of fear of childbirth, nor CB-PTSD or PPD symptoms, at 6 months postpartum.^84^

#### Mother-Infant-Focused Interventions

Mother-infant dyad interventions target the maternal-infant interaction through various modalities including skin-to-skin contact and play sessions. Improvement in the mother-infant interaction is thought to promote maternal mental health.^85–87^ Dyad interventions were tested in 3 RCTs including 1 secondary prevention and 2 tertiary preventions.^88–90^ Study quality scores were Good (23-26).

Immediate postpartum mother-infant skin-to-skin contact can have positive effects in reducing CB-PTSD symptoms following traumatic childbirth.^91^ Skin-to-skin during the ‘magical’ first postpartum hour vs. TAU (routine postpartum skin-to-skin) in 84 women meeting DSM PTSD Criterion A was associated with fewer CB-PTSD symptoms 2 weeks and 3 months post-intervention.^88^

Brief therapist-led one-on-one consecutive dyad observational and play intervention sessions following prematurity and performed at a later postpartum time point show positive outcomes in improving maternal sensitivity and post-traumatic stress symptoms. A 3-phase (33 and 42 weeks post-conception, and infant age 4 months) intervention of ∼5 observational and free play sessions (several hours in total) of mothers and their premature infants improved the quality of interactions in comparison with TAU (preterm without intervention) in a randomized sub-sample of 26 pairs.^89^ The treatment was associated with increase in maternal sensitivity and infant cooperation, decrease in infant difficulty, and significant decrease in CB-PTSD symptoms from time of intervention up to 12 months postpartum.^89^ In contrast, an earlier dyad-focused intervention initiated ∼33 days postpartum and during infant NICU hospitalization stay was not associated with improved outcomes. A series of 6 sessions (5 in NICU and the last at home at 2-4 weeks post-discharge) focused on reading infants’ cues and responding was not found more helpful than TAU (standard care) in 121 women with very low birth weight infants.^90^

#### Other Psychologically Oriented Interventions

Other psychological interventions for CB-PTSD include biofeedback tested as primary prevention in an NRCT;^92^ and a cognitive visuospatial task tested as secondary and tertiary preventions in an RCT and NRCT, respectively.^93, 94^ Study quality score ranged from Fair to Good (18-26).

Visual biofeedback ultrasound during the second stage of labor, involving the physician conveying a visual representation for the future mother of her pushing efforts and fetus movement in real time, shows benefits in reducing CB-PTSD risk. In an NRCT of 95 nulliparous women,^92^ ∼5 minutes of biofeedback vs. TAU (standard obstetrical coaching) increased maternal-newborn connectedness in the immediate postpartum, which in turn was associated with reduced acute stress in initial postpartum days and subsequently reduced CB-PTSD symptoms at 1 month.^92^ There were no direct effects of the treatment on CB-PTSD symptoms.

A brief visuospatial cognitive task procedure performed in the immediate postpartum following emergency Cesarean delivery shows short-term positive effects.^93^ This therapy is thought to interfere with consolidation of the traumatic visual memory, making the memory less perceptual and less intrusive.^95–97^ A single 15-minute computer game Tetris session within 6 hours postpartum vs. TAU in a randomized sample of 56 women was found acceptable by subjects and was associated with fewer intrusive traumatic memories of childbirth 1 week post-delivery.^93^ However, no significant treatment effects were observed for CB-PTSD, anxiety, or depression symptoms at 1 month postpartum.^93^ Likewise, in a pilot NRCT of 18 women (without control group) with severe childbirth-related re-experiencing symptoms ∼2 years postpartum, administered a single 20-minute Tetris session during childbirth recollection for the purpose of traumatic memory blockage,^94^ the majority reported fewer intrusive memories 1-2 weeks and 5-6 weeks post-intervention. For subjects who met CB-PTSD diagnosis, none met diagnosis at 1 month post-intervention.

#### Educational Interventions

Antenatal education aims to help expecting mothers via strategies for managing pregnancy, childbirth, and parenthood, and may also include postpartum interventions.^98–100^ Education interventions are provided by midwives and nurses. This review included 1 RCT^101^ and 1 NRCT^102^ primary educational prevention and 1 RCT secondary educational prevention.^103^ Study quality scores ranged from Good to Excellent (22-31).

Antenatal educational consecutive group sessions show benefit in non-high-risk women.^102^ Consecutive (4, 240 minutes) sessions focused on psychological and physiological adaption vs. TAU in a non-randomized sample of 90 second- and third-trimester pregnant women were associated with less fear of childbirth in pregnancy and more expected self-efficacy, and later at 6-8 weeks postpartum, with less fear of childbirth and fewer CB-PTSD symptoms.^102^ Likewise, consecutive one-on-one sessions focused on developing a birth plan vs. TAU in a randomized sample of 106 non-high-risk third-trimester women were associated with less fear of childbirth, improved childbirth experience, and fewer CB-PTSD and PPD symptoms 4-6 weeks post-delivery.^101^

In contrast, an early postpartum educational intervention utilizing self-help materials for women who had traumatic childbirth and were at risk for CB-PTSD without professional support was insufficient to reduce CB-PTSD symptoms. In an RCT of 678 women meeting PTSD DSM-IV Criterion A, subjects receiving self-help materials on how to manage early psychological responses during postpartum visit plus usual care, vs. TAU, did not show reduction in incidence of CB-PTSD diagnosis or sub-diagnosis assessed 6-12 weeks postpartum.^103^

## Comment

### a. Principal Findings

This systematic review provides insight derived from published randomized and non-randomized clinical trials of interventions tested in pregnant and postpartum women to inform evidence-based recommendations for primary and secondary prevention of CB-PTSD, and guidance for determining treatment approaches. Available studies (N=33) reviewed here range in quality between Poor and Excellent. They demonstrate that structured trauma-focused therapies and semi-structured midwife-led psychological counseling strategies are promising treatments (Figure 2). Other treatments to consider are traumatic memory blockage, mother-infant dyadic focused, and educational interventions (Figure 2). As additional RCTs are conducted, stronger evidence to support the efficacy of treatments for primary, secondary, or tertiary approaches will become available.

**Figure 2.**
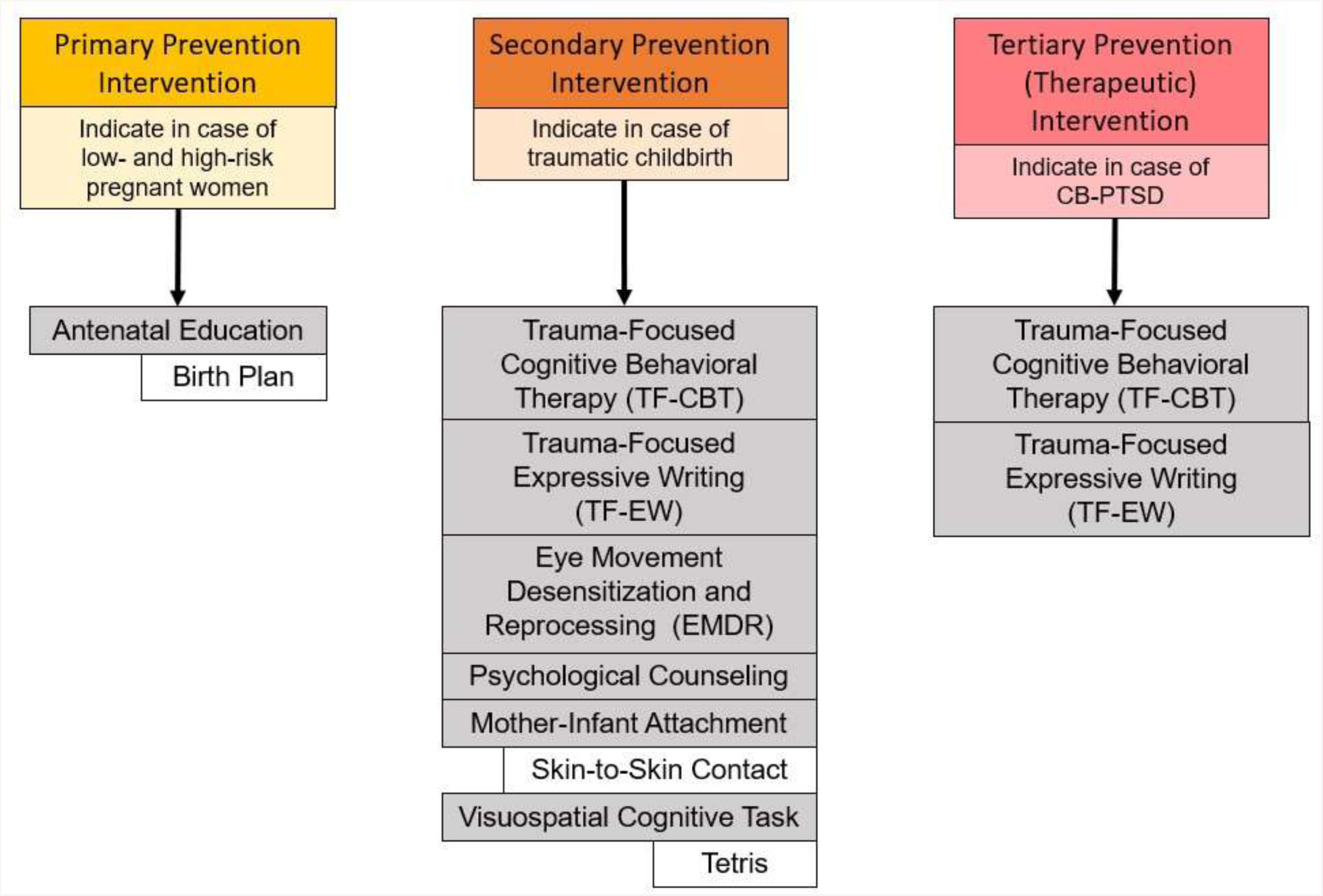
Recommended primary, secondary, and tertiary (i.e., therapeutic) interventions to mitigate or prevent the development of childbirth-related post-traumatic stress disorder (CB-PTSD). Recommendations are based on 16 studies employing randomized controlled clinical trials (RCTs) and reporting positive results. Grey boxes indicate categories of therapy strategy, and white boxes indicate specific implementations of those strategies. Trauma-Focused Expressive Writing (TF-EW) for secondary prevention was tested in universal samples.

An array of brief postpartum psychological interventions are safe, acceptable, and feasible to implement as early treatment, often before CB-PTSD presents as a clinically diagnosable disorder, thus minimizing serious consequences. A total of 16 RCTs reveal positive outcomes (Figure 2). Among them, the secondary preventions appear promising for reducing CB-PTSD symptoms compared with usual care in women exposed to traumatic childbirth. Evidence also supports the potential positive sustained effects of brief therapies (1-4 sessions) performed within 48-96 hours postpartum and during maternity hospitalization stay. This “in-house” approach could greatly facilitate access to postpartum care. Although psychological debriefing following childbirth trauma may not be helpful,^40, 53^ the few available RCTs suggest the effectiveness of EMDR^67^ and Trauma-Focused Expressive Writing (TF-EW),^72–75^ which largely target fear extinction through reprocessing of the trauma memory; one or few sessions of psychological counseling led by a midwife near bedside;^80–83^ and interventions focused on the mother-infant dyad (and skin-to-skin contact)^88–90^ during the “sensitive period” following childbirth. This latter approach suggests a second therapeutic target. What remains unclear is whether useful interventions delivered in the early postpartum have efficacy as standalone treatments for women with acute clinically significant traumatic stress and whether they can reduce CB-PTSD diagnosis incidence.

Antepartum educational interventions delivered universally to pregnant women before childbirth may promote positive mental outcomes during pregnancy and following childbirth.^101, 102^ The limited available evidence, based on two studies, suggests that universal interventions focused on birth plan and preparation are helpful, regardless of potential for exposure to traumatic childbirth. Postpartum educational interventions targeting women experiencing traumatic childbirth do not appear sufficient for reducing CB-PTSD incidence,^103^ underscoring the importance of the timing of educational interventions.

Interventions for the indication of CB-PTSD (tertiary prevention) with the goals of preventing worsening symptoms and improving functioning for women who endorse symptoms or have a diagnosis may have substantial benefits for the developing child. This review identified 5 RCT-tested interventions supporting the potential benefits of trauma-focused therapies (expressive writing and TF-CBT).

### b. Comparison with Existing Literature

A large body of literature addresses treatment approaches for PTSD in non-postpartum individuals.^104–106^ Although trauma-focused interventions are the gold standard, they suffer from high dropout rate,^107–109^ and some individuals with PTSD will remain treatment resistant.^110–112^ This underscores the importance of intervening effectively in the aftermath of trauma to buffer the development of persistent symptoms.

Currently, limited data are available on effective interventions to prevent PTSD.^45–47^ Childbirth, however, provides a unique opportunity to test early post-birth therapies for PTSD stemming from traumatic childbirth, facilitated by immediate access to postpartum patients. This review provides new insight on promising secondary preventive approaches for CB-PTSD, including the benefits of intervening in the very first post-trauma exposure days, which, with more replicated and high-quality studies, could inform clinical recommendations. This review expands the emerging literature on CB-PTSD therapies by covering trials published through December 2022. The available data favor targeted rather than universal approaches to treat postpartum women.

### c. Strengths and Limitations

This review adopts a comprehensive approach to evaluate available data on preventive interventions and treatments for CB-PTSD via quality assessment of all clinical trials published to date, not limited to a specific treatment modality, treatment time period, or maternal population. Hence, we provide insight into all three types of potential interventions, what the optimal timing for intervention may be, and which populations will benefit most. We use a well-validated standardized quantitative approach based on the PRISMA guidelines^49^ for study selection and data extraction, and assess external validity, internal validity, and power, to evaluate the published trials’ quality. While the primary outcome is CB-PTSD, we also consider co-morbid conditions, such as postpartum depression. Nevertheless, several limitations are worth noting. This review’s quality assessment was performed for each treatment modality separately, and grouping RCTs and non-RCTs studies into the relevant category. The main limitation in this approach is the small number of trials in some categories, which may limit the interpretation of the quality score range. Some studies lacked information about sample characteristics, degree of pre-treatment CB-PTSD severity, and clear time point of treatment outcome assessment, and these characteristics are only partly reflected in the assessment scale. Likewise, the definition of high-risk women exposed to childbirth trauma varied among studies and may have affected the ability to detect treatment effects. We did not intend to perform meta-analyses, which may have provided additional information. Finally, the number of published trials per prevention type is limited, which may prevent drawing strong conclusions.

### d. Conclusions and Implications

Maternal psychiatric morbidities are a leading complication of childbirth^113–115^ and involve heavy public health costs.^116–118^ Substantial evidence shows that a significant portion of women experiencing traumatic childbirth develop persistent symptoms of childbirth-related PTSD (CB-PTSD),^3, 12, 13, 119^ which cause functional impairment.^24^ Standards are lacking regarding what type of psychological therapy should be routinely delivered in postpartum care for the prevention or indication of this disorder, and this can have adverse consequences far beyond the directly affected postpartum woman. The available studies covered in this review suggest that intervening early in the postpartum period, and as soon as feasible, may reduce trauma reactions and in turn prevent CB-PTSD diagnosis. As a first step, this would require accurate identification of high-risk women who have experienced complicated, traumatic childbirth and may also show clinically significant acute stress.^120^ Early therapy delivered to high-risk women, rather than universally in the maternal population, would allocate available resources to those most in need and lower medical costs. A second critical step is ensuring treatment uptake during postpartum care. As presented in this review, manualized brief therapies delivered during maternity hospitalization stay offer a promising time window for effective therapy that also has the advantage of improving equity in care. A primary preventive approach in high-risk women may involve interventions focused on preparation for forthcoming childbirth delivered to pregnant women when they are already in frequent contact with health providers and during a time of motivation for self-care.

Important areas for future research include replicating the reported studies using adequate sample sizes, assessing long-term outcomes in RCT designs, shifting from exclusive patient self-report to also including mechanistic biomarkers, and identifying the golden hours following childbirth to maximize treatment response and uptake. Additionally, testing adjunctive or alternative non-trauma-focused intervention approaches that appear promising in individuals with general PTSD (e.g., mindfulness,^121^ yoga,^122^ metacognitive therapy (MCT)),^123^ and regarding resilience and psychological growth,^124^ therapies to enhance those traits, as well as the use of safe drug therapies (e.g., intra-nasal oxytocin),^125^ will expand available treatment options.

Ultimately, a personalized treatment approach incorporating therapeutic acceptability to the pregnant or postpartum woman and considering degree of symptom severity rather than a “one size fits all” strategy is likely to maximize treatment effectiveness. Based on the current state of knowledge, perinatal and mental health providers are strongly encouraged to consider on a case-by-case basis promising treatment options to prevent post-traumatic stress in the wake of childbirth trauma.

***Appendix A:*** Modified Downs and Black Checklist used in this systematic review.

## Data Availability

Table 1 of this manuscript presents a summary of the key findings of this systematic review.

