## Supplementary material for "A Systematic Review of Interventions for Prevention and Treatment of Post-Traumatic Stress Disorder Following Childbirth": Table 1

**Table 1.** ***Descriptions of clinical trials of interventions for the prevention or treatment of childbirth-related maternal PTSD (CB-PTSD)***

| First Author (Year) | Country | Study Design | Sample Size | Sample Characteristics | Sample Type | Preventative or Treatment | Treatment Modality | Intervention | Outcome Tim Points | Results | Outcome Measures | Quality Score |
|---|---|---|---|---|---|---|---|---|---|---|---|---|
| Selkirk (2006)[55] | AUS | RCT | N=149 | Not specified | Universal | 2˚ | Debriefing (P) | 1 session within 72h PP<br>Control: TAU | T1: ≤48hrs PP<br>T2: 1mo PP<br>T3: 3mo PP | No significant Tx effect on PPD, anxiety, and parenting distress. Tx was associated with more negative traumatic birth perceptions following OB interventions. Although Tx was associated with better dyadic satisfaction and was acceptable | CB-PTSD (IES); PPD (EPDS); Anxiety (STAI); Parenting stress (PSI); Perception of the birth (POBS); Dyadic satisfaction (DAS); Feedback of debriefing (FAD) | 19 |
| Priest (2003)[56] | AUS | RCT | N=174 | Not specified | Universal | 2˚ | Debriefing, CIS (P) | 1 session within 72hr PP<br>Control: TAU | T1: 2mo PP<br>T2: 6mo PP<br>T3: 12mo PP | No significant Tx effect on CB-PTSD and PPD symptoms, although the majority of the women reported finding the Tx helpful | CB-PTSD (IES); PPD (EPDS); Psychological Interview (SADS*) | 28 |
| Kershaw (2005)[58] | GBR | RCT | N=319 | 85% White<br>91% Partnered<br>Majority employed | Targeted | 2˚ | Debriefing, CIS (P) | 2 sessions at 10 days and 10wks PP<br>Control: TAU | T1: 10 days PP<br>T2: 10wks PP<br>T3: 20wks PP | No positive Tx effect on CB-PTSD symptoms or FoC | CB-PTSD (IES); FoC (WDEQ) | 26 |
| Abdollahpour (2019)[57] | IRN | RCT | N=193 | Age, M = 26 yrs | Targeted | 2˚ | Debriefing or CBC (P) | 1 debriefing or CBC session within 48hrs PP<br>Control: TAU | T1: 4-6wks PP<br>T2: 3mo PP | Positive debriefing Tx effect on CB-PTSD symptoms at T1. Positive CBC Tx effect on CB-PTSD symptoms at T1 and T2 | CB-PTSD (IES-R) | 22 |
| Meades (2011)[59] | GBR | NRCT | N=80 | Age, M = 34 yrs | Targeted | T | Debriefing (P) | 1 session within 1.3-72.2mo PP<br>Control: TAU. | T1: 1mo post Intervention | Positive Tx effect on CB-PTSD and negative appraisals of childbirth. No positive Tx effect on PPD | CB-PTSD (PSS-SR); PPD (EPDS); Perceived support (SOS); Negative appraisal (PTCI) | 17 |
| Jotzo (2005)[60] | DEU | NRCT | N=50 | Age, M = 31yrs<br>98% Partnered | Targeted | 2˚ | Trauma-preventative Crisis Intervention Program (P) | 1 session within 5 days PP with availabilities of psychological support<br>Control: TAU with availability of pastoral support. | T1: Post intervention | Positive Tx effect on overall CB-PTSD symptoms and as well as intrusion, avoidance, hyperarousal | CB-PTSD (IES); Peritraumatic dissociative experiences (PDEQ) | 19 |
| Shaw (2013,2014)[62,63] | USA | RCT | N=105 | Age, M = 32yrs<br>61% White<br>96% Partnered<br>53% Income ≥$100K | Targeted | 2˚ | CBT-TF (P) | 6 sessions within 1-2wks PP<br>Control: TAU with 1 informational session.<br>6-mo follow-up to Shaw 2013, with the option of three additional CBT sessions (total 9 sessions) | T1: 1wk post intervention<br>T2: 4-5wks PP<br>T3: 6mo PP | Positive Tx effect on CB-PTSD and PPD symptoms. Anxiety symptoms in both the control and intervention groups alleviated.<br>Tx effect remained at T3 only for women who received additional sessions | CB-PTSD (TES; DTS); Traumatic events (PSS:NICU; MINI-I*);Anxiety (BAI); PPD (BDI-II); Stress (SASRQ); PSS:NICU; Illness health Severity Index | 27 |

| Author Year | Country | Study Design | Sample Size | Sample Characteristics | Sample Type | Preventative or Treatment | Treatment Modality | Intervention | Outcome Time Points | Results | Outcome Measures | Quality Score |
|---|---|---|---|---|---|---|---|---|---|---|---|---|
| Simon (2021)[64] | USA | NRCT | N=19 | 32% White<br>100% Partnered<br>89% Employed<br>68% ≥ Bachelor's degree | Targeted | 2˚ | Trauma-focused group intervention (P) | 6 sessions over 3wks following preterm birth (exact PP timeframe not specified) | T1: 6wks post-intervention<br>T2: 6mo PP | Positive Tx effect on PPD at T1 and CB-PTSD and anxiety at T2 | CB-PTSD (DTS); Anxiety (BAI); PPD (BDI-II); Maternal Satisfaction Questionnaire; Fidelity Rating Scales | 20 |
| Nieminen (2016)[66] | SWE | RCT | N=56 | Age, M = 35yrs<br>95% Partnered<br>68% Employed<br>80% ≥ University degree | Targeted | T | iCBT-TF (P) | 8 sessions over 8-wks within ~2.8 years PP<br>Delayed control: Tx 5mo later. | T1: 8wks post-intervention<br>T2: Post-intervention for delayed Tx group (control) | Positive Tx effect on CB-PTSD, PPD and anxiety in Tx and delayed Tx groups. Positive Tx effect on quality of life in Tx group at T1 | CB-PTSD (MINI-I*; TES; IES-R); PPD (BDI-II; PHQ-9); Anxiety (BAI); Quality of Life (QOLI; EQ5D). | 26 |
| Sjömark (2022)[65] | SWE | RCT | N=266 | Age, M = 32yrs<br>98% Partnered<br>73% University degree | Targeted | T | iCBT-TF (P) | Part I: 6-wk online-based protocol ~2-4mo PP<br>Part II: structured weekly therapeutic support, email-based program given over 8-wks after Part I<br>Control: TAU | T1: 6wks post-intervention<br>T2: 14wks post-intervention<br>T3: 1yr post-intervention | There was no direct Tx effect on CB-PTSD at any time point between groups. Over time, both the control and intervention group experienced similar recovery trajectories and beneficial outcomes | CB-PTSD (TES); PPD (EPDS); Quality of Life (SWLS); Coping abilities (WCQ). | 23 |
| Chiorino (2020)[67] | ITA | RCT | N=37 | Age, M = 34yrs<br>86% Italian<br>97% Partnered<br>62% Employed<br>60% ≥ University degree | Targeted | 2˚ | EMDR (P) | 1 session within 3 days PP<br>Control: TAU with a single session of psychological supportive therapy | T1: 6wks PP<br>T2: 3mo PP | Positive Tx effect on CB-PTSD symptoms, flashbacks, and distress at T1. Fewer symptoms reported at T2 in Tx group compared to TAU, however, no significant between group differences observed. No positive Tx effects on mother-infant-bonding or PPD | CB-PTSD (IES-R; PDEQ); PPD (EPDS); Mother-Infant-Bonding (MIBS) | 25 |
| Sandström (2008)[68] | SWE | Pilot NRCT | N=4 | Age, M = 28yrs<br>100% Partnered | Targeted | T | EMDR (P) | Multiple sessions in women with previous traumatic births | T1: Post intervention<br>T2: 1-3yrs post intervention | Positive Tx effect on reported CB-PTSD symptom reduction at T1, 75% positive Tx effect at T2 and reports of improved infant-bonding among 2 patients | CB-PTSD (TES); Data from interviews and psychotherapist notes* | 13 |
| Kranenburg (2022)[69] | NLD | NRCT | N=26 | Age, M=32 | Targeted | T | EMDR (P) | Up to 8 (mean 5) weekly sessions delivered ≥4-wks PP (mean 10-mo) in women with CB-PTSD or severe PTSD-symptoms and a comorbid mental health condition | T1: Post-intervention, time not specified | Positive Tx effect on reported CB-PTSD symptoms and among patients who met criteria for CB-PTSD diagnosis at study inclusion, all participants lost their CB-PTSD diagnoses post-intervention | CB-PTSD (PCL-5, LEC-5); Childhood Trauma (CTQ); Childbirth Perception (CPS). | 19 |

| Author Year | Country | Study Design | Sample Size | Sample Characteristics | Sample Type | Preventative or Treatment | Treatment Modality | Intervention | Outcome Time Points | Results | Outcome Measures | Quality Score |
|---|---|---|---|---|---|---|---|---|---|---|---|---|
| Di Blasio (2002)[72] | ITA | RCT | N=64 | Age, M = 33yrs<br>33% ≥ High School | Universal | 2˚ | EW (P) | 1 session 2 days PP writing about thought/feelings of delivery experience<br>Control: TAU, no writing task | T1: 48hrs PP<br>T2: 2mo PP | Positive between group Tx effect on CB-PTSD hyperarousal and avoidance symptoms at T1; between-group avoidance and re-experiencing symptoms, but not hyperarousal, were also significantly different at T2 | CB-PTSD (PPQ) | 21 |
| Di Blasio (2009)[73] | ITA | RCT | N=242 | Age, M = 32yrs<br>100% Partnered<br>89% ≥ high school | Universal | 2˚ | EW (P) | 1 session 2 days PP writing about thought/feelings of delivery experience<br>Control: TAU, no writing task | T1: 48hrs PP<br>T2: 2mo PP<br>T3: 12mo PP | Positive Tx effect maintained on CB-PTSD symptoms and on hyperarousal and avoidance symptoms at T1, no effect on intrusive symptoms.<br>At T3, all symptoms decreased except for hyperarousal | CB-PTSD (PPQ) | 22 |
| Di Blasio (2015)[74] | ITA | RCT | N=176 | Age, M = 32yrs<br>84% Partnered<br>78% Employed<br>85% ≥ High School | Universal | 2˚ | EW (P) | 1 session within 96hrs PP writing about thought/feelings of delivery experience<br>Control: writing about daily events | T1: 96hrs PP<br>T2: 3mo PP | Positive Tx effect on PPD symptoms (T1,T2) and on CB-PTSD symptoms (T2) | CB-PTSD (LASC; PPQ); PPD (BDI-II) | 23 |
| Di Blasio (2015)[75] | ITA | RCT | N=113 | Age, M = 31 yrs<br>80% Partnered<br>80% Employed<br>86% ≥ High School | Universal | 2˚ | EW (P) | 2 sessions within 96hrs PP writing about thought/feelings of delivery experience<br>Control: writing about daily events | T1: 3mo PP | Positive Tx effect on CB-PTSD and PPD symptoms at T1, especially for women with high-to-moderate PTS symptoms | CB-PTSD (PPQ); PPD (BDI-II); Essay Content | 20 |
| Horsch (2016)[76] | CHE | RCT | N=67 | Age, M = 31yrs<br>56% Partnered | Targeted | T | EW (P) | 3 writing tasks in 3 consecutive days at 3mo PP<br>Control: TAU | T1: 4mo PP<br>T2: 6mo PP | Positive Tx effect on CB-PTSD and PPD symptoms and overall improved mental health status at both time points | CB-PTSD (PPQ); PPD (EPDS); Mental and physical health status (SF-36); Scores of diseases (CRIB II; the Perinatal risk Inventory); Reports on intervention satisfaction. | 27 |
| Barry (2001)[77] | USA | RCT | N=38 | Age, M = 33yrs<br>89% Partnered | Targeted | T | EW (P) | 4 writing tasks in 4 consecutive days within 2-14 mo of infant NICU hospitalization<br>Control: Waiting-list control | T1: Post-intervention | Positive Tx effect on CB-PTSD symptoms | CB-PTSD (SCL-90-R; IES-R). | 21 |
| Asadzadeh (2020)[81] | IRN | RCT | N=90 | Age, M = 25yrs<br>40% University degree | Targeted | 2˚ | Counseling (P) | 1 session within 72hrs of delivery, 1 telephone session at 4-6wks PP<br>Control: TAU | T1: 4-6wks PP<br>T2: 3mo PP | Positive Tx effect on CB-PTSD, PPD, and anxiety (T1 and T2). Similar treatment effects were observed in the control group at T2, although the decrease in symptoms was not as pronounced | CB-PTSD Checklist (PCL-5); PPD (EPDS); Anxiety (HAM-A*) | 24 |

| Author Year | Country | Study Design | Sample Size | Sample Characteristics | Sample Type | Preventative or Treatment | Treatment Modality | Intervention | Outcome Time Points | Results | Outcome Measures | Quality Score |
|---|---|---|---|---|---|---|---|---|---|---|---|---|
| Gamble (2005)[82] | AUS | RCT | N=103 | Age, M = 28yrs<br>93% White<br>85% Partnered<br>60% ≥ High school | Targeted | 2˚ | Counseling (P) | 1 in-person session within 72hrs of delivery + 1 telephone session at 4-6wks PP<br>Control: TAU | T1: 4-6wks PP<br>T2: 3mo PP | Positive Tx effect CB-PTSD, PPD, feelings of self-blame and confidence about a future pregnancy | CB-PTSD (MINI-I*); PPD (EPDS; DASS-21); Anxiety/Stress (DASS-21); Self-blame and confidence about a future pregnancy (MSSS). | 23 |
| Ryding (1998)[83] | SWE | RCT | N=99 | Age, M = 30 yrs | Targeted | 2˚ | Counseling (P) | 3 or 4 sessions delivered during the first days to 3wks PP<br>Control: TAU | T1: 1mo PP<br>T2: 6mo PP | Positive Tx effect on CB-PTSD and cognitive appraisal at T1-T2. No positive Tx effect on women with the most serious stress reactions | CB-PTSD (IES); Cognitive appraisal of delivery and distress (W-DEQ; SCL) | 18 |
| Bahari (2022)[80] | IRN | RCT | N=166 | Age, M = 27yrs<br>11% Employed<br>63% ≥ high school | Targeted | 2˚ | Counseling (P) | 2 in-person sessions at 24-48hrs and 10-15 days PP<br>1 telephone session 4-6wks PP<br>Control: TAU | T1: 2mo PP | Positive Tx effect in alleviating severity of PTSD symptoms and PPD in addition to improving mother-bonding | CB-PTSD (PCL-5); PPD (EPDS); Mother-Infant Bonding (PBQ) | 26 |
| Ryding (2004)[84] | SWE | RCT | N=162 | Age, M = 32 yrs | Targeted | T | Group Counseling (P) | 2 group sessions at 1-2mo PP<br>Control: TAU | T1: 6mo PP | No positive Tx effect on CB-PTSD, PPD, or FoC | CB-PTSD (IES); PPD (EPDS); FoC (WDEQ) | 23 |
| Abdollahpour (2016)[88] | IRN | RCT | N=84 | Age, M = 26 yrs | Targeted | 2˚ | Skin-to-skin Contact (B) | SSC according with 9-instinctive stage magical hour protocol in the immediate post-birth period<br>Control: TAU | T1: 2wks PP<br>T2: 4-6wks PP<br>T3: 3mo PP | Positive Tx effect on CB-PTSD at T1 and T3, but not T2 | CB-PTSD (IES-R) | 23 |
| Borghini (2014)[89] | CHE | RCT | N=26 | Age, M = 32 yrs | Targeted | T | Mother-Infant Attachment (P) | Infant observation at 33wks PP, clinical interview at 42wks PP, 3 sessions at 4mo PP, 1wk apart, of mother-infant free play<br>Control: (1) TAU in mothers of preterm infants without intervention, and (2) of mothers with term infants | T1: 42wks PP<br>T2: 4mo PP<br>T3: 12mo PP | Positive Tx effect on CB-PTSD at all time points and increased maternal sensitivity and infant cooperation at T2 in the preterm with intervention group | PTS (PPQ); Quality of mother-child interactions (NBAS) | 26 |
| Zelkowitz (2011)[90] | CAN | RCT | N=121 | Age, M = 31yrs<br>88% Partnered<br>6% Employed<br>66% ≥ Junior college<br>Income, M = $59K | Targeted | T | Cognitive Intervention and Mother-Infant Attachment (Cues group) (P) | 6 sessions focused on teaching mothers how to cope and interact with their children ~33d ±12d PP to 6-8wks PP<br>Control: TAU, standard infant care resources | T1: Immediate post-intervention timeframe | No positive Tx effect on levels of CB-PTSD, PPD, and anxiety | CB-PTSD (PPQ); PPD (EPDS); Anxiety (STAI); Stress (PSI) | 23 |

| Author Year | Country | Study Design | Sample Size | Sample Characteristics | Sample Type | Preventative or Treatment | Treatment Modality | Intervention | Outcome Time Points | Results | Outcome Measures | Quality Score |
|---|---|---|---|---|---|---|---|---|---|---|---|---|
| Schlesinger (2022)[92] | ISR | NRCT | N=95 | Age, M = 29yrs<br>91% Partnered<br>79% Employed | Universal | 1° | Visual Biofeedback (P) | 1 session during labor<br>Control: TAU, standard labor | T1: 2d PP<br>T2: 1mo PP | Positive Tx effect on CB-PTSD symptoms via increased feelings of maternal connectedness, which was associated with reduced acute stress levels at T1. There was an indirect Tx effect between visual biofeedback and decreased CB-PTSD symptoms at T2 | CB-PTSD (PLC-S); Stress (SASRQ); Fear of Childbirth (PTS-FC); Maternal attachment self-report Likert-type scale | 18 |
| Horsch (2017)[93] | CHE | RCT | N=56 | Age, M = 33yrs<br>91% White<br>88% Partnered<br>64% ≥ Bachelor's degree | Targeted | 2° | Visuospatial Cognitive Task (P) | 1 Tetris session within 6hrs of delivery after an ECS<br>Control: TAU | T1: 1wk PP<br>T2: 1mo PP | Positive Tx effect on self-reported acute stress symptoms and reduction in the number of intrusive memories at T1. Significant group differences at T2 on PTSD criteria and PSD avoidance symptom | CB-PTSD (PDS); Anxiety and PPD (HADS); Distress (ASDS); Diary accounts of intrusive memories | 26 |
| Deforges (2022)[94] | CHE | NRCT | N=18 | Age, M=33<br>94% Partnered<br>56% University degree | Targeted | T | Visuospatial Cognitive Task (P) | 1 Tetris session delivered >7mo PP | T1: 1-2wks post-intervention<br>T2: 1mo post-intervention | Greater than 50% reduction in CB-PTSD intrusive memories at T1 and T2. CB-PTSD symptom severity significantly reduced at T2 for most women (58.8%). Of the 8 women previously diagnosed with CB-PTSD, none met diagnostic criteria post-intervention | CB-PTSD (PCL-5); Diary report of intrusive memories | 23 |
| Gökçe (2016)[102] | TUR | NRCT | N=90 | Age, M = 26yrs<br>61% ≥ Bachelor's degree | Universal | 1° | Antenatal education (E) | 4 weekly group sessions<br>Control: TAU | T1: 6-8wks PP | Positive Tx effect on CB-PTSD, FoC, and childbirth self-efficacy | CB-PTSD (IES-R); FoC (WDEQ); Childbirth Self-efficacy Inventory | 22 |
| Slade (2020)[103] | UK | RCT | N=678 | Age, M = 30yrs<br>89% White<br>89% Partnered | Targeted | 2° | Educational self-help strategies (E) | Self-help materials to go through after delivery<br>Control: TAU | T1: 6-12wks PP | No positive Tx effect on CB-PTSD reduction in CB-PTSD incidence rates | CB-PTSD (CAPS-5* interview); Depression and anxiety (HADS); Attachment (MPAS); Couple relationship quality (DAS4) | 31 |

| Author Year | Country | Study Design | Sample Size | Sample Characteristics | Sample Type | Preventative or Treatment | Treatment Modality | Intervention | Outcome Time Points | Results | Outcome Measures | Quality Score |
|---|---|---|---|---|---|---|---|---|---|---|---|---|

| | | | | | | | | | | | | |
|---|---|---|---|---|---|---|---|---|---|---|---|---|
| Ahmadpour (2022)[101] | IRN | RCT | N=106 | Age, M=26yrs<br>13% University degree | Universal | 1˚ | Birth Plan (E) | 2 sessions.<br>Control: TAU | T1: Immediate PP<br>T2: 4-6wks PP | Positive Tx effect on CB-PTSD symptom reduction, improved childbirth experience, less fear of delivery, and lower EPDS in the intervention group compared to control | CB-PTSD (PSS)<br>PPD (EPDS)<br>FoC (W-DEQ)<br>Childbirth Experience (CEQ 2.0; DFS ; SCIB) | 26 |

**Quality Scores:** Excellent (29-31); Good (22-28); Fair (17-21); Poor (≤16).

**Abbreviations:**

* = Clinician-Administered Assessment

Intervention Categories:
1˚: Primary
2˚: Secondary

Intervention Types:
(B): Behavioral Intervention
(E): Educational Intervention
(P): Psychological Intervention

Country Abbreviations:
TUR: Turkey
AUS: Australia
USA: United States of America
IRN: Iran
SWE: Sweden
DEU: Germany
ITA: Italy
UK: United Kingdom
CAN: Canada
CHE: Switzerland
ISR: Israel

Additional Abbreviations:
ASDS: Acute Stress Disorder Scale
BAI: Beck Anxiety Inventory
BDI-II: Beck Depression Inventory-II
CBC: Cognitive Behavioral Counseling
CB-PTSD: Childbirth-related Post-Traumatic Stress Disorder
CBT(-TF): Cognitive Behavioral Therapy (Trauma-Focused)
CIS-Debriefing: Critical Incidence Stress Debriefing
CRIB II: Clinical Risk Index for Babies
CSE: Collective Self-Esteem Scale
DTS: Davidson Trauma Scale
EMDR: Eye Movement Desensitization & Processing
EPDS: Edinburgh Postnatal Depression Scale
EPL: Everyday Problems List
EQ5D: EuroQol 5 Dimensions
EW: Expressive Writing
FoC: Fear of Childbirth
HAM-A: Hamilton Anxiety Rating Scale
iCBT: Internet-based CBT
IES(-R): Impact of Events Scale (Revised)
LASC: Los Angeles Symptoms Checklist
M: Mean
MFI: Multidimensional Fatigue Inventory

MIBS: Mother to Infant Bonding Scale
MINI-I: Mini International Neuropsychiatric Interview
MSSS: Modified Social Support Survey
NBAS: Neonatal Behavioral Assessment Scale
NICU: Neonatal Intensive Care Unit
NRCT: Non-Randomized Controlled Trial
PBQ: Postpartum Bonding Questionnaire
PCL-5: PTSD Checklist for DSM-5
PERI: Perinatal Risk Inventory
PDEQ : Peritraumatic Dissociative Experiences Questionnaire
PHQ-9: Patient Health Questionnaire
POBS: Perception of Birth Scale
PP: Postpartum
PPD: Postpartum Depression
PPQ: Perinatal Posttraumatic Stress Disorder Questionnaire
PSI: Parenting Stress Index
PSS: Parental Stressor Scale
PSS-SR: PTSD Symptom Scale – Self-Report Version
PTCI: Posttraumatic Cognitions Scale
QOLI: Quality Of Life Inventory
QPI: Quality of provider interactions
RCT: Randomized Controlled Trial
SADS-1: Schedule for Affective Disorders & Schizophrenia
SCL: Symptoms Checklist
SF-36: 36-Item Short Form Health Survey questionnaire
SOS: Significant Others Scale
SSC: Skin to Skin Contact
STAI: State-Trait Anxiety Inventory
T: Treatment
TES: Traumatic Event Scale
TES-B: Traumatic Event Scale – Delivery
WDEQ: Wijma Delivery Expectancy Questionnaire
